# Comparison of tobacco index versus smoking status in the prevalence of elevated blood pressure levels

**DOI:** 10.1101/2024.05.31.24308301

**Authors:** Víctor Juan Vera-Ponce, Fiorella E. Zuzunaga-Montoya, Lupita Ana Maria Valladolid-Sandoval, Luisa Erika Milagros Vásquez-Romero, Joan A. Loayza-Castro, Stella M. Chenet, Felix Llanos Tejada

## Abstract

**Introduction:** Smoking is a well-established risk factor for hypertension (HTN) and high blood pressure (HBP); however, there are controversies regarding whether it is merely the act of smoking that increases the risk of HTN or if it is the quantity of tobacco consumed that influences it.

**Objective:** To determine the relationship between different smoking statuses and the tobacco index (GTI) with the presence of HBP and HTN in the Peruvian population.

**Methods:** An analytical cross-sectional study based on data obtained from Peru’s Demographic and Family Health Survey (ENDES) from 2014-2022. The primary variable in this study is blood pressure level, which was classified into normotension, HBP, and HTN. Smoking status was categorized into smoker status (never, ex, current, daily smoker), daily smoker (yes versus no), and the GTI.

**Results:** Overall, daily smokers showed a higher prevalence of HTN, while those with a TI ≥ 20 showed a higher prevalence of both HBP and HTN. Significant discrepancies were observed regarding sex and the presence of these diseases, although GTI remained a consistent associated factor.

**Conclusions:** The tobacco index is highlighted as a more reliable indicator of the risk associated with smoking compared to simply categorizing individuals as smokers or non-smokers. This index, which quantifies both the frequency and duration of smoking habits, has proven to be a robust factor in the prevalence of these pathologies.

## Introduction

Smoking is a significant risk factor for chronic disease. According to the WHO report, tobacco use globally causes about 8 million deaths every year, with many of these being caused by second-hand smoke exposure ^(1)^. Not only does smoking impact health, but it also affects thousands of people’s quality of life ^(2)^.

Despite global efforts to reduce tobacco smoking, prevalence rates in some countries remain high. Specific populations continue to exhibit alarming levels of increase, impeding progress against this significant public health issue. However, according to the Demographic and Family Health Survey (ENDES) results, we observe mixed signals when focusing specifically on the Peruvian population. The survey indicated that the percentage of smokers decreased, averaging just over 6% in 2020, before slightly increasing to nearly 7% ^(3)^.

Smoking is a well-known risk factor for both Hypertension (HTN) and High Blood Pressure (HBP), as supported by numerous studies ^(4–6)^. However, some reports have suggested that in addition to being smokers themselves, the total amount of tobacco consumed could also contribute to HTN. Research has shown variation among smokers in terms of developing HTN, based on findings from various sources ^(7–10)^, indicating the need to understand how quantity affects the development of HTN. This highlights the importance of further research exploring the relationship between different groups’ smoking status, such as never-smoked individuals, former smokers, current smokers, and other categories considered for the Tobacco Index (GTI), and the impact on factors such as HBP and diseases like HTD.

## Methods

### Type and Design of Research

This analytical cross-sectional study is based on data from the ENDES of Peru, collected between 2014 and 2022. The ENDES is a national survey conducted annually in Peru, designed to provide representative data on various demographic and health aspects at the national, regional, and departmental levels. The ENDES is carried out by the National Institute of Statistics and Informatics (INEI) and uses a probabilistic, multi-stage, and stratified sampling design to ensure the representativeness of its results.

### Population, Sample, and Eligibility Criteria

The population consists of individuals who participated in the ENDES during 2014-2022. For this study, the sample will include participants aged 18 years or older with systolic blood pressure (SBP) and diastolic blood pressure (DBP) measured during the survey. Participants who reported a prior diagnosis of HTN will be excluded from the analysis, as awareness of this condition could have altered their smoking behavior, introducing bias in the results. Additionally, individuals whose blood pressure values are implausible or outside the physiologically acceptable range will be excluded to ensure the accuracy and validity of the data used in the analysis ^(11)^.

### Variables and Measurement

The primary variable in this study is blood pressure status. To define this variable, two states will be considered ^(12)^ :

- The first is HBP, an SBP between 130- and 139 mm Hg or a DBP between 80 and 89 mm Hg.
- The second is HTN, defined as an SBP greater than or equal to 140 mm Hg and a DBP greater than or equal to 90 mm Hg.

The primary dependent variable was smoking status, which was classified in three distinct ways:

- First, smoking status, grouping individuals into four categories: never smoker (individuals who have never smoked), former smoker (individuals who have smoked in the past but do not currently smoke), current non-daily smoker (individuals who currently smoke but not every day), and daily smoker (individuals who smoke every day) ^(13)^.
- Second, the daily smoker variable distinguishes between those who smoke daily and those who do not (including those who do not smoke at all and those who do not smoke daily).
- Finally, the GTI, calculated based on the number of cigarettes smoked daily and the number of years the individual has smoked, is categorized into three groups: 0, 1 to 19, and 20 or more. This index allows for the evaluation of not only whether a person smokes but also the total exposure to tobacco smoke over time ^(14)^.

Several covariables will be included to adjust the analyses and consider possible confounding factors. These covariables are sex, classified as male and female; age group, divided into four categories: 20-35 years, 36-59 years, 60-69 years, and 70 years or older; and marital status, distinguishing between being in a partnership (married or cohabiting) and being single (with and without a partner). Additionally, the region of residence (Metropolitan Lima, other coastal areas, highlands, and jungle), wealth index (classified into quintiles from poorest to richest), and educational level (no schooling or primary versus secondary or higher) will be included. Furthermore, area of residence, classified as urban or rural; nutritional status, categorized as normal weight, overweight, and obesity according to body mass index (BMI); alcohol consumption, distinguishing between non-consumption or non-excessive consumption and excessive consumption; and altitude levels will be considered.

#### Procedures

A standardized process was implemented to ensure consistent results when reliably and accurately measuring systolic and diastolic blood pressure levels. Participants’ blood pressure levels were recorded using a digital sphygmomanometer (OMRON, model HEM-713). Two types of cuffs were used, adapted to everyone’s arm circumference: standard cuff (220-320 mm) and large cuff (330-430 mm).

Measurements were carefully taken in a controlled environment, ensuring participants were comfortable and seated with their right arm extended on a heart-level surface. The first measurement was taken after a 5-minute relaxation interval, and the second was taken two minutes after the first; this interval between the two measurements allowed cardiovascular parameters to stabilize. Subsequently, the mean of everyone’s systolic and diastolic blood pressure readings was calculated, and this average value was used as the basis for subsequent analyses. This procedure provides clinicians with a more reliable means of assessment by stabilizing blood pressure fluctuations and capturing a more accurate picture of a person’s typical blood pressure levels.

The smoking status variable was evaluated through self-reporting by participants. A series of specific questions were used to classify subjects into different smoking categories. These questions included: “In the past 12 months, have you smoked cigarettes?”, “In the past 30 days, have you smoked cigarettes?” “Do you smoke daily?” “Age you started smoking daily - Unit,” “Age you started smoking daily,” “How many years ago did you start smoking daily (in years),” and “How many cigarettes do you smoke daily?”.

In addition to collecting the data above, the ENDES also gathered relevant sociodemographic information necessary for informing the research. For more details on how the evaluations were conducted, it is recommended to visit the report published annually on the INEI website ^(15)^

#### Statistical Analysis

Statistical analyses were performed using R software version 3.14. Variables were summarized in absolute and relative frequencies for a detailed sample description. Subsequently, a multinomial logistic regression analysis was conducted, given that the blood pressure status variable has the three previously mentioned categories.

Three distinct multinomial logistic regression analyses assessed the association between smoking status and blood pressure states. The first analysis focused on smoking status, daily smoking, and the tobacco index. The results of these analyses were presented as adjusted odds ratios (aOR) for the covariates mentioned earlier; additionally, p-values and 95% confidence intervals (CI) were reported for each association measure presented.

#### Ethical Considerations

The database used in this study, ENDES, is publicly accessible and available for unrestricted use. Both initial and follow-up evaluation data are accessible to the general public. The survey information is anonymous, lacks personal identifiers, and is collected without direct contact with human subjects (14). Therefore, since the data used are secondary and do not involve interaction with individuals or collecting new personal information, submitting this study for ethical committee review was not considered necessary.

## Results

Regarding the general characteristics of the study sample (n = 231,834), 49.41% were women. Most participants (62.29%) were between 20 and 44 years old, 23.18% were between 45 and 59, and 14.52% were 60 or older. Regarding educational level, 22.94% had primary education, 42.09% had secondary education, and 34.72% had higher education. Regarding marital status, 33.88% were single, and 66.12% lived with a partner. The geographical distribution showed that 33.76% resided in Metropolitan Lima and 76.28% in rural areas.

Concerning smoking status, 8.37% were former smokers, 9.83% currently smoked, and 1.70% were daily smokers. Specifically evaluating daily smoking, it was found that 1.70% of the participants smoked daily. Regarding the GTI, 1.52% had an index between 0 and 20, and 0.07% had an index of 20 or more.

Regarding lifestyle, only 2.76% consumed alcohol excessively, and 8.77% consumed at least five servings of fruits/vegetables daily. In terms of BMI, 41.71% were obese. Additionally, 97.25% did not have diabetes. Regarding the altitude of residence, 70.80% lived at less than 1500 meters. Finally, the prevalence of HBP and HTN was 14.73% and 12.44%, respectively. The rest of the characteristics can be seen in Table 1.

**Table 1.** Demographic characteristics of participants

| **Characteristic** | **n = 231,834** |
| --- | --- |
| **Sex** |  |
| Female | 114,543 (49.41%) |
| Male | 117,292 (50.59%) |
| **Group age** |  |
| 20 - 44 years | 144,420 (62.29%) |
| 45 - 59 years | 53,746 (23.18%) |
| 60 to more | 33,668 (14.52%) |
| **Educational Level** |  |
| No Level | 458 (0.25%) |
| Primary | 42,180 (22.94%) |
| Secondary | 77,386 (42.09%) |
| Superior | 63,826 (34.72%) |
| **Civil status** |  |
| Single | 61,302 (33.88%) |
| With a partner | 119,627 (66.12%) |
| **Natural region** |  |
| Metropolitan Lima | 78,268 (33.76%) |
| Resy of coast | 59,278 (25.57%) |
| Mountain Range | 63,888 (27.56%) |
| Jungle | 30,401 (13.11%) |
| **Area of residence** |  |
| Rural | 176,848 (76.28%) |
| Urbano | 54,986 (23.72%) |
| **Wealth index** |  |
| The poorest/Poor | 95,698 (41.28%) |
| Medium | 47,170 (20.35%) |
| Rich/Richest | 88,966.88 (38.38%) |
| **Smoker Status** |  |
| Never | 185,697 (80.10%) |
| Former Smoker | 19,408 (8.37%) |
| Currently Smoker | 22,787 (9.83%) |
| Daily Smoker | 3,942 (1.70%) |
| **Smoker Dially** |  |
| No | 227,892 (98.30%) |
| Yes | 3,942 (1.70%) |
| **Global Tobacco Index** |  |
| Cero | 228,130 (98.40%) |
| >= 0 or < 20 | 3,531 (1.52%) |
| >= 20 | 174 (0.07%) |
| **Alcohol consumption** |  |
| Never or non-excessive alcohol consumption | 225,442 (97.24%) |
| Excessive alcohol consumption | 6,392 (2.76%) |
| **Fruit and vegetable consumption** |  |
| **≥ 5 servings per day** |  |
| No | 211,505 (91.23%) |
| Yes | 20,329 (8.77%) |
| **Body mass index** |  |
| Normal weight | 79,887 (34.47%) |
| Overweight | 55,217 (23.82%) |
| Obesity | 96,687 (41.71%) |
| **T2DM** |  |
| No | 225,349 (97.25%) |
| Yes | 6,365 (2.75%) |
| **Altitude** |  |
| 0 to 1499 | 164,092 (70.80%) |
| 1500 to 2499 | 15,736 (6.79%) |
| 2500 to 3499 | 32,726 (14.12%) |
| 3500 or more | 19,226 (8.29%) |
| **Pressure Level** |  |
| Normotension | 168,862 (72.84%) |
| High blood pressure | 34,141 (14.73%) |
| Hypertension | 28,831 (12.44%) |

**Table 2.** Adjusted Multinomial Logistic Regression Analysis for the Association between Smoking Status and Blood Pressure Levels

| Characteristic | High blood pressure |  | Hypertension |  |
| --- | --- | --- | --- | --- |
|  | aOR* | 95% CI | aOR* | 95% CI |
| **General** |  |  |  |  |
| **Smoker Status** |  |  |  |  |
| Never | Ref. |  | Ref. |  |
| Former Smoker | 0.97 | 0.93, 1.01 | **0.84** | **0.80, 0.89** |
| Currently Smoker | **1.06** | **1.01, 1.11** | 0.99 | 0.94, 1.04 |
| Daily Smoker | 1.04 | 0.95, 1.14 | 1.06 | 0.96, 1.16 |
| **Smoker Dially** |  |  |  |  |
| No | Ref. |  | Ref. |  |
| Yes | 1.07 | 0.96, 1.18 | **1.11** | **1.11, 1.23** |
| **Global Tobacco Index** |  |  |  |  |
| Cero | Ref. |  | Ref. |  |
| ≥ 0 or < 20 | 1.01 | 0.92, 1.11 | 1.05 | 0.95, 1.16 |
| ≥ 20 | **1.84** | **1.19, 2.84** | **1.76** | **1.16, 2.68** |
| **Women** |  |  |  |  |
| **Smoker Status** |  |  |  |  |
| Never | Ref. |  | Ref. |  |
| Former Smoker | 0.86 | 0.73, 1.02 | **0.71** | **0.58, 0.88** |
| Currently Smoker | **1.38** | **1.20, 1.58** | 0.98 | 0.82, 1.17 |
| Daily Smoker | 0.96 | 0.71, 1.29 | 1.28 | 0.96, 1.69 |
| **Smoker Dially** |  |  |  |  |
| No | Ref. |  | Ref. |  |
| Yes | **0.57** | **0.40, 0.81** | 1.18 | 0.83, 1.66 |
| **Global Tobacco Index** |  |  |  |  |
| Cero | Ref. |  | Ref. |  |
| ≥ 0 or < 20 | 0.84 | 0.60, 1.17 | **1.41** | **1.05, 1.89** |
| ≥ 20 | **3.28** | **1.28, 8.37** | 0.38 | 0.07, 2.09 |
| **Men** |  |  |  |  |
| **Smoker Status** |  |  |  |  |
| Never | Ref. |  | Ref. |  |
| Former Smoker | **0.93** | **0.89, 0.98** | **0.82** | **0.78, 0.87** |
| Currently Smoker | 0.99 | 0.95, 1.03 | 0.95 | 0.90, 1.00 |
| Daily Smoker | 1.04 | 0.95, 1.15 | 1.04 | 0.93, 1.15 |
| **Smoker Dially** |  |  |  |  |
| No | Ref. |  | Ref. |  |
| Yes | 1.07 | 0.97, 1.18 | 1.11 | 0.99, 1.24 |
| **Global Tobacco Index** |  |  |  |  |
| Cero | Ref. |  | Ref. |  |
| ≥ 0 or < 20 | 1.04 | 0.94, 1.14 | 1.03 | 0.93, 1.15 |
| ≥ 20 | **1.74** | **1.07, 2.81** | **2.16** | **1.38, 3.36** |

\*Each smoking status variable has been independently adjusted for sex, group age, educational level, civil status, natural region, area of residence, wealth index, alcohol consumption, nutritional status, diabetes, altitude, and consumption of fruits and vegetables.
aRP: Adjusted Prevalence ratio. 95% CI: 95% confidence interval
Source: self-made

According to the adjusted multivariable analysis, several significant associations were found between smoking status and blood pressure levels. Generally, former smokers exhibited a protective effect against HTN (aOR: 0.84; 95% CI 0.80, 0.89), whereas current smokers showed a higher prevalence of HBP (aOR: 1.06; 95% CI 1.01, 1.11). Daily smokers also had a higher probability of presenting HTN (aOR: 1.11; 95% CI 1.11, 1.23). Meanwhile, those with a TI of ≥ 20 showed a higher prevalence of HBP (aOR: 1.84; 95% CI 1.19, 2.84) and HTN (aOR: 1.76; 95% CI 1.16, 2.68).

In women, current smokers were observed to have a higher probability of presenting HBP (aOR: 1.38; 95% CI 1.20, 1.58), while former smokers exhibited a protective effect against HTN (aOR: 0.71; 95% CI 0.58, 0.88). Daily smokers showed a protective effect against HBP (aOR: 0.57; 95% CI 0.40, 0.81). Regarding the GTI, those with an index of ≥ 20 had a higher probability of presenting HBP (aOR: 3.28; 95% CI 1.28, 8.37), and those with an index ≥ 0 and < 20 presented a higher risk of HTN (aOR: 1.41; 95% CI 1.05, 1.89).

In men, former smokers exhibited a protective effect against HBP (aOR: 0.93; 95% CI 0.89, 0.98) and HTN (aOR: 0.82; 95% CI 0.78, 0.87). A GTI of ≥ 20 was associated with a higher prevalence of HBP (aOR: 1.74; 95% CI 1.07, 2.81) and HTN (aOR: 2.16; 95% CI 1.38, 3.36).

## Discussion

In the present study, discrepancies were found in the results, as the probabilities of having HBP or HTN vary depending on how smoking status is evaluated and specifically by sex. This finding suggests that it is essential to ask whether a person smokes and how much they smoke.

### Comparison with other studies

The importance of quantifying not only the presence of smoking but also its intensity has been supported by multiple studies. In our study, the observed discrepancies in the results reflect this complexity and highlight that the frequency and the amount of tobacco consumed are critical to understanding the impacts on cardiovascular health. For example, the study by Nance et al. ^(9)^ indicated that merely evaluating smoking status (as a former smoker or current smoker) is insufficient to assess the risk of cardiovascular diseases, finding that while duration was not associated with cardiovascular diseases, current intensity was, thus using pack-years would be an appropriate measure.

On the other hand, the study by Kim et al. ^(16)^ found significant associations between smoking and the risk of type II diabetes in men but not in women. For men, both early initiation of smoking and the cumulative amount of tobacco smoked (measured in pack-years) were associated with a higher risk of developing type II diabetes. This highlights the importance of using pack years as a risk measure for metabolic diseases.

Additionally, the article by Fan and Zhang ^(7)^ suggests that it is not simply the act of smoking or not smoking that influences the risk of hypertension but the intensity and accumulation of the smoking habit. The study found that men with light or moderate smoking trajectories and those with high cumulative tobacco exposure (measured in pack-years) had a higher risk of developing hypertension compared to non-smokers. This indicates that both the number of cigarettes smoked daily and the duration of the smoking habit are essential factors in increasing the risk of hypertension.

Our findings further suggest that former smokers experience a protective effect against HTN in both men and women. This effect could be due to significant lifestyle changes after quitting smoking, such as adopting a healthier diet and increasing physical activity^(17,18)^. This phenomenon reflects the resilience and recovery capacity of the human body from the damage caused by smoking and highlights the importance of supporting smokers in the cessation process.

Additionally, we observed significant differences between sexes in response to smoking. Current smokers show a higher likelihood of HBP in women, while former smokers exhibit a protective effect against HTN. In men, both HBP and HTN are linked to current smoking and a high tobacco index. Other studies have found these sex differences in the risk of metabolic diseases ^(7,9,16)^. These variations suggest that the effects of smoking on cardiovascular health may differ substantially between sexes, underscoring the need for personalized and targeted intervention strategies that consider these differences ^(19)^.

Conversely, the finding that daily female smokers have a lower risk of HBP is surprising and suggests several reasons. One could be biased in the question regarding daily smoking activity in this group. Another could be the existence of a subgroup of women possibly more resilient to the negative impact of tobacco or the influence of unmeasured confounding factors in this study ^(7,9,16)^. These findings highlight the complexity of the risk patterns associated with smoking and the need for more sophisticated analytical approaches to unravel these effects.

The TI, which measures total accumulated exposure to tobacco, showed a strong association with the prevalence of HBP and HTN, especially in those with an index of ≥ 20, more so in men. This underscores the importance of considering the total tobacco burden when assessing cardiovascular risk ^(20–23)^. This quantitative approach to tobacco consumption could significantly improve the accuracy of our risk predictive models and, consequently, prevention and treatment strategies.

#### Contribution to the Field

This study sheds light on public health by showing how smoking impacts heart health in men and women based on how much they smoke. Not just the act of smoking but also the quantity smoked plays a significant role in disease rates. Hence, it is vital for public health policies to push not only quitting but also cutting down on cigarettes for those who cannot quit entirely. This can lead to better intervention and teaching plans that cater to various groups within society.

Moreover, this research hints that strategies to manage smoking impact on blood pressure should vary by sex due to different responses observed. Tailoring these methods can help craft more personalized and efficient interventions; these would target both preventing high blood pressure (HTN) and fostering a healthy lifestyle among ex-smokers. Knowing the benefits of quitting, along with gender differences in risk, can help shape more vital public messages and cessation campaigns aimed at specific groups effectively.

This study highlights the importance of broader tobacco use measures in judging heart risk by showing how the TI can predict HTN. Such findings back expanding smoke watch programs that not only count smoking rates but also track changes in habits over time; these measures are crucial for assessing how well public programs on cutting down smoking work now and in the future.

#### Study Limitations

However, some limitations of this study must be noted when interpreting its results. First off, using self-reported data for smoker status might bring recall or social desirability bias—this could skew classifying participants as smokers or smokers. Plus, since the study uses a cross-sectional design, it prevents proven cause-and-effect relationships between smoking and blood pressure levels; hence, we avoid discerning if smoking leads to hypertension or if it is the other way around. Moreover, even though many confounding factors were adjusted for, there might still be unmeasured ones distorting our observed relationships. These limitations point out why longitudinal studies and more objective ways of measuring are needed to evaluate how smoking impacts heart health accurately.

## Conclusions

This study highlights the tobacco index as a more reliable indicator of the risk associated with smoking compared to simply categorizing individuals as smokers or non-smokers. This index, which quantifies both the frequency and duration of the smoking habit, has proven to be a robust predictor of the prevalence of elevated blood pressure levels.

Moreover, our analysis revealed significant sex differences in the relationship between the tobacco index and blood pressure levels. Women and men appear to respond differently to smoking in terms of its impact on blood pressure, underscoring the need for differentiated approaches in public health interventions and future research. This sex variability could guide the personalization of smoking cessation interventions, ensuring they are more effective and relevant for each group.

Given the importance of the tobacco index in evaluating cardiovascular risk and the observed sex differences, we recommend the implementation of national surveys specifically designed to assess smoking habits. These surveys should be designed to capture not only whether individuals smoke but also how much and for how long they have smoked, allowing for more accurate estimates of the tobacco index. Such data could provide a more substantial basis for the development of public health policies, prevention programs, and intervention strategies focused not only on smoking cessation but also on reducing consumption among active smokers.

## Data Availability

The data supporting the findings of this study can be accessed from the original research paper at the following link: https://proyectos.inei.gob.pe/microdatos/

## Acknowledgments

A special thanks to the members of the Tropical Diseases Research Institute, Universidad Nacional Toribio Rodríguez de Mendoza de Amazonas (UNTRM), Amazonas, Peru, for their support and contributions throughout the completion of this research.

## Informed Consent

It was not necessary to obtain informed consent in this study.

## Author Contributions

**Víctor Juan Vera-Ponce:** Conceptualization, Investigation, Methodology, Resources, Writing - Original Draft, Writing - Review & Editing

**Fiorella E. Zuzunaga-Montoya:** Conceptualization, Investigation, Methodology, Writing - Original Draft, Writing - Review & Editing

**Lupita Ana Maria Valladolid-Sandoval:** Investigation, Project Administration, Writing - Original Draft, Writing - Review & Editing

**Luisa Erika Milagros Vásquez-Romero:** Investigation, Resources, Writing - Original Draft, Writing - Review & Editing

**Joan A. Loayza-Castro:** Software, Data Curation, Formal Analysis, Writing - Review & Editing

**Stella M. Chenet:** Validation, Visualization, Writing - Original Draft, Writing - Review & Editing

**Felix Llanos Tejada:** Methodology, Supervision, Funding Acquisition, Writing - Review & Editing

